# ACE-2-like enzymatic activity is associated with immunoglobulin in COVID-19 patients

**DOI:** 10.1101/2022.12.19.22283685

**Authors:** Yufeng Song, Regan Myers, Frances Mehl, Lila Murphy, Bailey Brooks, Jeffrey M. Wilson, Alexandra Kadl, Judith Woodfolk, Steven L. Zeichner

## Abstract

Many mechanisms responsible for COVID-19 pathogenesis are well-established, but COVID-19 includes features with unclear pathogenesis, such as autonomic dysregulation, coagulopathies, and high levels of inflammation. The receptor for SARS-CoV-2 spike protein’s receptor binding domain (RBD) is angiotensin converting enzyme 2 (ACE2). We hypothesized that some COVID-19 patients may develop antibodies that have negative molecular image of RBD sufficiently similar to ACE2 to yield ACE2-like catalytic activity - ACE2-like abzymes. To explore this hypothesis, we studied patients hospitalized with COVID-19 who had plasma samples available obtained about 7 days after admission. ACE2 is a metalloprotease that requires Zn^2+^ for activity. However, we found that the plasma from some patients studied could specifically cleave a synthetic ACE2 peptide substrate, even though the plasma samples were collected using disodium ethylenediaminetetraacetate (EDTA) anticoagulant. When we spiked plasma with synthetic ACE2, no ACE2 substrate cleavage activity was observed unless Zn^2+^ was added or the plasma was diluted to decrease EDTA concentration. After processing samples by 100 kDa size exclusion columns and protein A/G adsorption, which depleted immunoglobulin by >99.99%, the plasma samples did not cleave the ACE2 substrate peptide. The data suggest that some patients with COVID-19 develop antibodies with abzyme-like activity capable of cleaving synthetic ACE2 substrate. Since abzymes can exhibit promiscuous substrate specificities compared to the enzyme whose active site image they resemble, and since proteolytic cascades regulate many physiologic processes, anti-RBD abzymes may contribute to some otherwise obscure COVID-19 pathogenesis.

## Introduction

The primary pathology in COVID-19 involves pulmonary disease^1–3^. Other end organ involvement, likely directly attributable to infection with the virus, can include gastrointestinal disease, such as diarrhea, and loss of smell and taste^2–4^. While the basic pathophysiology of COVID-19 is well-established, COVID-19 has many baffling features, including disorders involving high inflammation, disorders of the clotting cascade, and problems with blood pressure homeostasis^2, 5, 6^. Some sequelae do not develop until a week or more after infection, suggesting that their pathogenesis may result from secondary processes.

Reports suggest that some of the inflammatory processes associated with COVID-19 may relate to antibodies against the SARS-CoV-2 spike protein, S. For example, small numbers of children who have received COVID-19 vaccines and have had no evidence of infection with SARS-CoV-2, as well as a small number of infants born to mothers with COVID-19^7^ have been described as having multi-system inflammatory syndrome of children (MIS-C)^8^.

In addition to acute COVID-19, some patients experience Post-Acute Sequelae of SARS CoV-2 infection (PASC, sometimes called “long COVID”), a heterogeneous group of symptoms that continue after acute infection has resolved and can include evidence of a hyperinflammatory state^9^, persistent coagulopathies, and neurophysiologic dysregulation, for example aberrant blood pressure regulation. While some clinical features observable at the beginning of the COVID-19 clinical course, such as SARS-CoV-2 viremia, autoantibodies, inflammatory markers, and activation of Epstein-Barr Virus, may be associated with an increased risk of long COVID^10^, the detailed pathogenic processes responsible for PASC remain unclear.

Membrane-bound angiotensin converting enzyme 2 (ACE2) is the main cellular receptor for SARS-CoV-2. Binding of the SARS-CoV-2 spike protein receptor binding domain (RBD) to ACE2 initiates the pathway that leads to infection of a new host cell. ACE2, a Zn^2+^ metalloenzyme, cleaves angiotensin II into angiotensin 1-7, which has vasodilatory activity. Angiotensin converting enzyme (ACE) cleaves angiotensin I to yield angiotensin II, which has vasoconstrictive and hypertensive effects. ACE2 therefore helps to counteract the hypertensive effects of angiotensin II, and so has blood pressure counterregulatory activity compared to ACE. There have been isolated reports and small case series describing persistent hypotension in patients with COVID-19 and patients with PASC without a clearly identified cause^11–13^.

The kallikrein-kinin system is a proteolytic cascade that helps regulate inflammation (reviewed in ^14^). There is cross-talk between the coagulation, kallikrein-kinin, complement and renin-angiotensin systems^15^. Some patients with severe COVID-19 respiratory disease have been found to have altered regulation of the kallikrein-kinin system in bronchoalveolar lavage fluid^16^. Abnormalities in the kallikrein-kinin system have been proposed as contributing to the pathogenesis of severe COVID-19^17^.

Catalytic antibodies, “abzymes”^18, 19^, are antibodies that have catalytic activity. When abzymes were first described, they generated considerable excitement. Investigators hypothesized that it would be possible to produce clinically and biotechnologically useful abzymes.

However, with additional studies, it became clearer that abzyme activity was significantly lower than the activity of more conventional enzymes (*k*cat/*Km* values ∼10^2^ - 10^4^ s^-1^•M s^-1^ ^20^ vs. *k*cat/*Km* values of ∼10^5^ s^-1^•M s^-1^)^21^. Hence, interest in the biotechnological applications of abzymes waned, but an appreciation of the potential role of abzymes in the pathogenesis of disease, including autoimmune disease, continued. High concentrations of abzymes in the circulation may compensate for the abzymes’ lower activity to yield clinically significant effects.

Patients with autoimmune diseases can make abzymes catalyzing, for example, cleavage of vasoactive intestinal peptide (VIP)^22^, DNA^23^, immunoglobulin components^24^, components of the clotting cascade^25–27^, and myelin basic protein (MBP) in patients with multiple sclerosis^28^. Abzymes that cleave the HIV envelope protein have been observed to occur in patients with HIV, so there is precedence for the induction of a catalytic antibody by a virus^29^.

Investigators have deliberately made anti-idiotypic abzymes with catalytic activity like the activity of a particular enzyme of interest^30, 31^. Investigators identify an antibody against an enzyme, and then produce an antibody against the anti-enzyme antibody. For some of the anti-idiotypic antibodies, there is enough resemblance to the active site of the enzyme that the anti-idiotypic antibody has catalytic activity resembling the enzyme^32, 33^. In some instances, the substrate specificity of the abzyme differs substantially from that of the original enzyme^31, 32, 34^, with the abzyme showing more substrate promiscuity than the original enzyme.

The receptor binding domain of the SARS-CoV-2 spike protein uses the ACE2 enzyme on the surface of the target future host cell as its receptor. The RBD is not an antibody, but it does bind ACE2. Taking as an example of abzyme production the anti-idiotypic antibody to an antibody against an enzyme, it is plausible to hypothesize that in some cases, an antibody against the RBD might resemble the ACE2 active site and so have catalytic activity. If an antibody directed against the RBD has some resemblance to ACE2, it is also plausible that anti-RBD antibodies, for some small fraction of patients, might have catalytic activity reminiscent of ACE2. We hypothesized that some COVID-19 patients may develop anti-RBD antibodies, with promiscuous catalytic activity that, in some cases, cleaves substrates in patients to yield clinically significant effects. FIG 1 outlines this hypothesis schematically. We therefore undertook a study to explore whether some COVID-19 patients might have antibodies that can cleave an ACE2 substrate.

**FIG 1.**
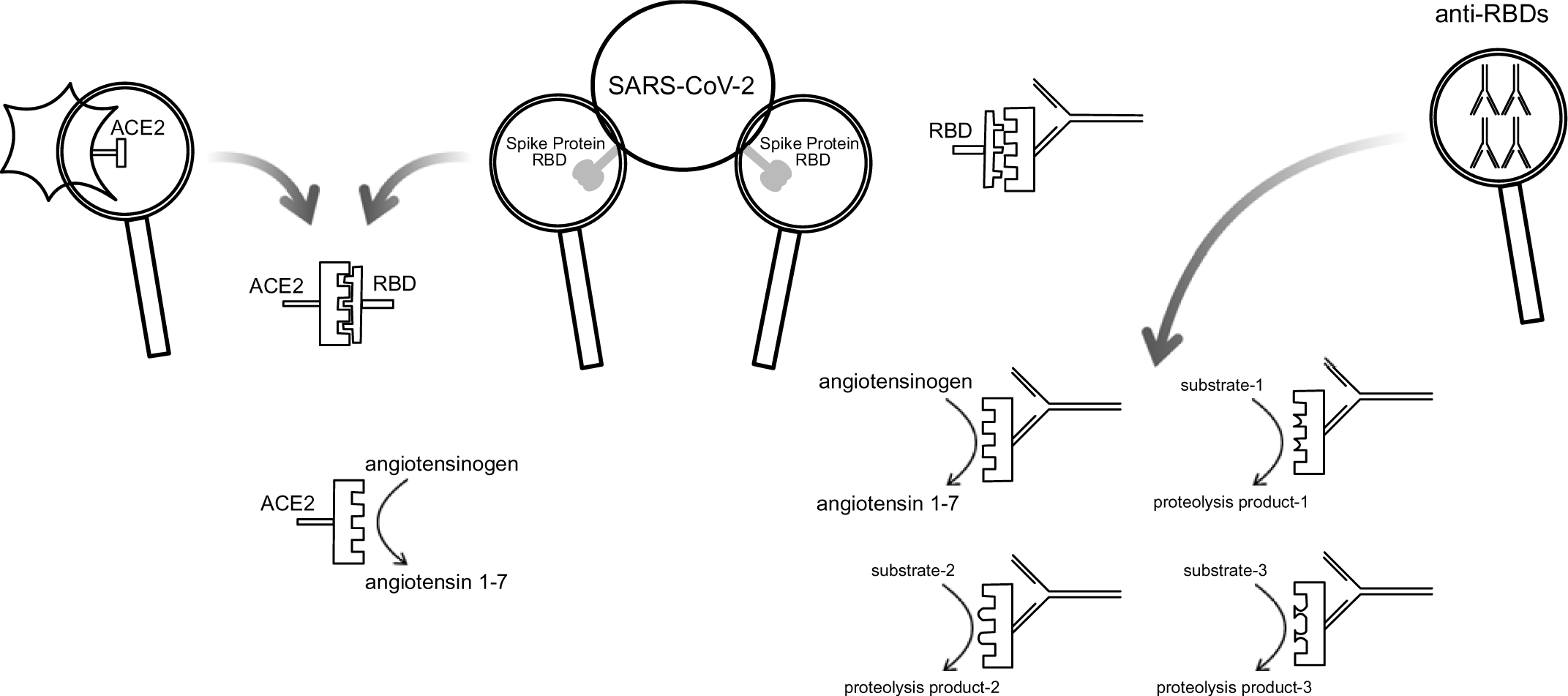
Schematic diagram illustrating the hypothesis underlying the study. The SARS-COV-2 Spike Protein Receptor Binding Domain (RBD) binds ACE2 on the surface of a potential host cells. The RBD has a partial negative image of ACE2. Some antibodies against the RBD may then have a conformation sufficiently like ACE2 to have proteolytic activity similar to the proteolytic activity of ACE2. The hypothesis further suggests that the catalytic activity of some antibodies will have a substrate specificity like ACE2, but because the negative image of RBD present antibodies is imperfect, some antibodies with proteolytic activity will have an altered substrate specificity, potentially affecting multiple proteolytic regulatory cascades.

We studied plasma samples from a group of 67 patients, with samples obtained relatively early in their disease course, on about the 7th day of hospitalization. As a negative control we purchased pooled human plasma that was collected in late 2018, about a year before the first cases of COVID-19 were described. We tested plasma using an established fluorometric assay for ACE2 activity, which detects cleavage of an ACE2 peptide substrate. We found that some of the patients had ACE2 substrate peptide specific cleavage activity when the assay was run directly on disodium ethylenediaminetetraacetate (EDTA)-anticoagulated plasma samples, even though ACE2 is a metalloprotease that requires Zn^2+^ for activity. Recombinant ACE2 added to plasma did not show ACE2 substrate-cleaving activity unless ZnCl_2_ was added to samples at concentrations greater than the EDTA. Processing the samples through a 100 KDa size exclusion column followed by absorption with protein A/G beads, which depleted IgG by >99.99%, eliminated the ability of the samples to cleave the ACE2 peptide substrate, supporting the hypothesis that a fraction of COVID-19 patients may develop antibodies that have ACE2 substrate cleavage activity.

## Methods

### Clinical Cohort

The clinical cohort was recently described in more detail in another publication^35^. The hypothesis motivating that work held that COVID-19 would lead to large scale reactivation of lymphotrophic herpesviruses EBV and HHV-6, but the data obtained in the study did not support that hypothesis. In brief, the University of Virginia (UVA) enrolled a prospective cohort of moderate to severe COVID-19 adult patients admitted to the hospital with a diagnosis confirmed by a nucleic acid amplification test. Patients consented were entered into the study. Clinical data were entered into study databases. Plasma was collected into clinical standard lavender top (EDTA anticoagulant) tubes. Samples were processed in the UVA Biorepository and Tissue Facility and stored at −80 C. Study participants (n=67) were adults ages 18 years and above. We used samples obtained on or about day 7 of hospitalization for this study, prior to anti-inflammatory drug administration (Table 1). The predominant viral strain for our sample was the alpha strain with 98.51% of our sample (66 patients) being collected before July of 2021.The sample collection protocol was approved by the UVA IRB (HSR #200110), and approval was obtained to work on the specimens (HSR #HSR200362), and it follows the Strengthening the Reporting of Observational Studies in Epidemiology (STROBE) guidelines. EDTA-anticoagulated healthy normal donor plasma was purchased from Valley Biomedical (http://www.valleybiomedical.com/), (Pooled Human Plasma, Cat. No. HP1051PK2, Lot No. 21M2548). This lot of plasma was collected on December 12, 2018 and December 13, 2018 (communication with Valley Biomedical technical support), about a year before the start of the COVID-19 pandemic, so it is extremely unlikely that the plasma was derived from any blood from patients infected with SARS-CoV-2. Plasma was aliquoted and stored at -20℃ according to manufacturer’s instructions.

**Table 1.** Patient Demographic and Clinical Characteristics (cohort previously described in ^35^) (IQR: interquartile range)

| **Characteristic** | **N = 67, median, (IQR)** |
| --- | --- |
| *Age at Admission* | 60 (48, 66) |
| *First Weight (kg)* | 91 (76, 112) |
| Unknown | 1 |
| *Height (m)* | 1.68 (1.60, 1.83) |
| Unknown | 6 |
| *Gender* |  |
| Female | 28 (42%) |
| Male | 39 (58%) |
| *Race* |  |
| African American | 23 (34%) |
| Native American | 1 (1.5%) |
| Caucasian | 31 (46%) |
| Other | 12 (18%) |
| *Smoking Status* |  |
| Current | 1 (2.1%) |
| Former | 14 (29%) |
| Never | 24 (50%) |
| Unknown | 9 (19%) |
| NA | 19 |
| *Need for ICU* |  |
| No | 28 (42%) |
| Yes | 39 (58%) |
| *Need for Intubation* |  |
| No | 41 (61%) |
| Yes | 26 (39%) |
| *Blood Type* |  |
| A- | 2 (4.8%) |
| A+ | 14 (33%) |
| AB+ | 1 (2.4%) |
| B+ | 3 (7.1%) |
| O- | 1 (2.4%) |
| O+ | 21 (50%) |
| Not determined | 25 |
| *Highest Temperature* |  |
| <38 | 42 (49%) |
| 101 | 12 (18%) |
| >38.5 | 12 (18%) |
| >40 | 1 (1.5%) |
| *Length of Stay (Days)* | 10 (5, 22) |
| *Time to Death (Days)* |  |
| 6 | 1 |
| 10-20 | 2 |
| 20-30 | 3 |
| >30 | 2 ( |
| *Highest D-Dimer* | 605 (280, 2,074) |
| *Highest CRP* | 10 (4, 16) |
| *BMI* | 33 (27, 38) |
| *Need for Vasopressors* |  |
| Yes | 44 (66%) |
| *Dexamethasone* |  |
| Yes | 41 (61%) |
| *Prednisone* |  |
| Yes | 3 (13%) |
| *Methylprednisolone* |  |
| Yes | 5 (22%) |
| *Tocilizumab* |  |
| Yes | 4 (6%) |
| *Remdesivir* |  |
| Yes | 27 (40%) |
| *Creatinine* | 1.00 (0.80, 1.45) |
| *Hgb* | 12.4 (10.8, 13.6) |
| *IFNa3* | 45 (36, 72) |
| Not determined | 45 |
| *Highest Ferritin* | 1277 (7, 7161) |
| Not determined | 5 |
| *Highest Troponin* |  |
| <0.04 | 47 (73%) |
| 0.04-0.4 | 12 (18%) |
| >0.6 | 3 (5%) |
| Not determined | 5 |
| *Highest Creatinine* | 1.10 (0.90, 1.80) |

### Angiotensin II Converting Enzyme (ACE2)-like Abzyme Activity Detection

Abzyme catalytic activities in plasma samples were measured using an Angiotensin II Converting Enzyme (ACE2) Activity Assay Kit, Fluorometric (Abcam, Cat. #ab273297) according to the manufacturer’s protocol. This assay employs a synthetic ACE2 peptide substrate labeled with a fluor and a quencher. The manufacturer’s product information claims a lower limit of detection for an unmodified assay of 400 µU. The manufacturer does not release detailed information concerning the peptide substrate, holding that information as a trade secret. The kit includes recombinant human ACE2 (ACE2) for use as a positive control. 50µl plasma was added to wells in a 96-well black flat bottom assay plate (Corning, Cat. #3603) and incubated at room temperature (RT), in the dark for 15 min. Then 50µl of substrate pre-diluted according to manufacturer’s protocol was added into the wells of the plate, mixing the substrate with the plasma sample. 25µM OMNIMMP® fluorogenic control substrate (Enzo Life Sciences, Cat. #MML-P127), Mca-Pro-Leu-OH which is suggested as fluorogenic Mca control peptide by manufacturer (https://www.enzolifesciences.com/BML-P163/mca-apk-dnp/), were added to the samples as ACE2 activity specific negative controls as mentioned previously ^36^. The plate was placed into a SpectraMax® M5 multi-model microplate reader (Molecular Devices) and the Relative Fluorescent Unit (RFU, Ex/Em = 320/420 nm) values were measured in kinetic mode every 20 min for a total of 16 hours at 37℃. ACE2 inhibitor provided in the assay kit was also used for additional controls.

### ACE2 Substrate Peptide Cleavage Activity in Plasma Samples in the Presence and Absence SARS-CoV-2 Spike Protein Synthetic Peptides

To evaluate the specificity of ACE2 substate cleavage activity by the plasma, we designed tiled SARS-CoV-2 spike protein RBD peptides which covered RBD amino acid sequence from residues Arg319 to Phe541 including receptor binding motif (RBM) from Ser438 to Gln506 based on previous comprehensive structure analysis on viral-host interaction ^37^, had them synthesized by SB-PEPTIDE (SmartBioscience SAS, France) (Table 2). Plasma samples and positive control, purified recombinant human ACE2 protein provided in the ACE2 assay kit (Abcam, Cat. #ab273297), were incubated with or without 5-fold serially diluted peptide pools at room temperature for 15min before adding the substrate. Samples incubated with ddH2O and DMSO which were equivalent to the concentrations used in the peptide pool reconstitution served as negative controls. Then 50 µL of ACE2 substrate (pre-diluted according to manufacturer’s protocol) was added into the wells and the samples were assayed. Relative Fluorescent Units were measured, and data was analyzed as described below.

**Table 2.**
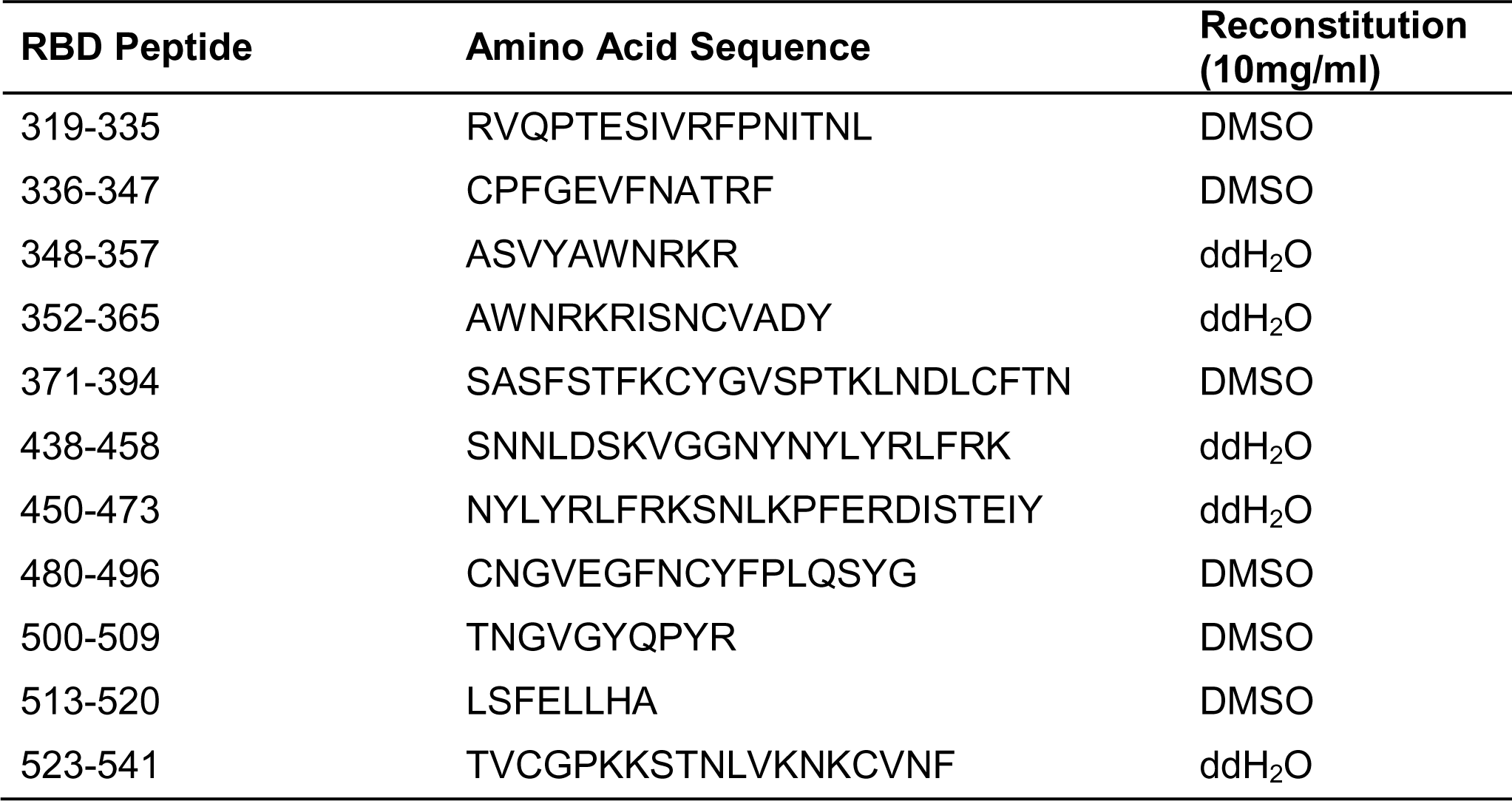
SARS-CoV-2 Spike RBD Peptides for ACE2-like Activity Competition Inhibition Assay.

### Determination of ACE2 Substrate Peptide Cleavage Activity in Plasma Samples With and Without Added ZnCl_2_ or Plasma Dilution

To evaluate whether native ACE2 shed into blood^38, 39^ could contribute to the catalytic activity observed in the patient plasma samples, we investigated whether EDTA present in the plasma samples inhibited activity of recombinant ACE2 spiked into the samples. EDTA can inhibit the catalytic activity of ACE2, a Zn^2+^ metaloprotease^40, 41^.

We conducted serial 10-fold dilutions of the normal donor plasma in ACE2 assay buffer from the assay kit and then added 2 µL recombinant positive control ACE2, provided as a positive control in the ACE2 assay kit (Abcam, Cat. #ab273297) to each well to the samples to the samples according to the manufacturer’s protocol, conducting the ACE2 assays on the serially diluted samples with the added recombinant ACE2. We also added ZnCl_2_ to some samples to achieve final concentrations ranging from 0 to 2 mM. Samples containing plasma, recombinant ACE2 and ZnCl_2_ mixture with pre-diluted ACE2 inhibitor served as negative comparisons. Then, 50 µL of substrate (pre-diluted according to manufacturer’s protocol) was added into the wells. Relative Fluorescent Units were measured, and data was analyzed as described below.

### Human Immunoglobulin Depletion and Detection

To deplete immunoglobulins from the samples, patient plasma samples or pooled healthy donor plasma were centrifuged at 12,000 g at 4 ℃ for 15 min and supernatants were passed through a 0.45 µm syringe filter (FisherScientific, Cat. #97204) to remove contaminating particulate matter. We then placed 500 µL of the filtered samples into ultrafiltration columns with 100kDa cut-off ultrafiltration membrane (Pierce™ Cat. # 88503) and centrifuged the columns at 12,000 × g at 4 ℃ until > 400 µL of filtrate was produced. To further deplete antibodies in the plasma samples, we then incubated the flow-through fractions with 30 µL pre-washed Protein A/G Magnetic Beads (Life Technologies, Cat. # 88803, 10 mg/mL) at room temperature (RT) for 1 hour, followed by magnetic removal of the beads, and repeated the same incubation procedure with another freshly prepared 30 µL magnetic beads. The resulting processed plasma samples were stored at 4℃ for short times (less than 12 hours) or -20 ℃ for longer times. Human IgG was measured using a Human IgG ELISA Kit (Abcam, Cat. #ab100547) according to the manufacturer’s protocol. We added 50 µL processed samples, as well as standards to multi-microwell strips and incubated them at room temperature for 2.5 hours. Strips were washed with wash buffer and 50 µL diluted biotin-labeled IgG detection antibody were then added to each well with subsequent HRP-streptavidin incubation after another wash step. Each incubation was performed at room temperature for at least 1 hour. After washing, 100 µL of TMB One-Step Substrate was added to each well and was incubated for 5 to 15 min at room temperature in the dark with gentle shanking. The reaction was stopped by adding 50 µL Stop Solution to each well. OD values were read at 450 nm using an accuSkan FC micro-well plate reader (ThermoFisher). IgG concentrations were calculated based on standard curves generated with the same batch of assay strips. Human IgM was measured using a Human IgM ELISA Kit (Invitrogen, Cat. #BMS2098) according to the manufacturer’s protocol. Briefly, after washing the strips twice, we added 100 µL processed samples, as well as standards together with 50 µL HRP-conjugated antibody to multi-microwell strips and incubated them at room temperature for at least 1 hour. Strips were washed with wash buffer. After washing, 100 µL of TMB One-Step Substrate was added to each well and was incubated for 5 to 15 min at room temperature in the dark with gentle shanking. The reaction was stopped by adding 100 µL Stop Solution to each well. OD values were read at 450 nm using an accuSkan FC micro-well plate reader (ThermoFisher). IgM concentrations were calculated based on standard curves generated with the same batch of assay strips.

### RBD Binding Assays

We measured IgG binding to the S-RBD in serum with a quantitative ImmunoCAP^TM^-based system, employing a Phadia 250 (Thermo-Fisher/Phadia), as described^42^. In brief, we biotinylated S-RBD (RayBiotech, Peachtree Corners, GA) and conjugated it to streptavidin-coated ImmunoCAP^TM^s (Thermo-Fisher/Phadia). We subtracted background signal by subtracting the signal due to unconjugated streptavidin ImmunoCAP^TM^, which was run in parallel with each sample.

### Data Analysis

Since the RFU values for the assays conducted on the samples reached 80% of maximum at about 240 min, RFU values observed over 240 min generated by an ACE2 kit were used to evaluate ACE2 substrate cleavage activity in the plasma samples. For each sample, a value corrected for baseline fluorescence in the sample was obtained by the subtracting the baseline RFU value from the value at each subsequent time and the sum of the baseline-corrected RFU values over 240 min was calculated using Excel. Normalized RFU in FIG 6 were calculated as following formulation:

> (**RFU***actual* - **RFU***minimun*)/(**RFU***maximal* - **RFU***minimum*)

For SARS-CoV-2 spike RBD peptide competition assay, the AUCs (area under curve) of each selected sample with individual incubation peptide pool amount were calculated based on all RFU readouts. Subsequent statistical analysis and data visualization was done using R (version 4.2.1) with the Rstudio environment and packages ggplot2, tidyverse, readr, drc, ggpubr, ggh4x, gridExtra, and scales. Samples with the top 20% and a control bottom 10% RFU values over 240 min were identified for antibody depletion and further assays.

## Results

To develop assays with enhanced sensitivity for ACE2 substrate cleavage activity, we used a commercially available kit, in which a synthetic peptide ACE2 substrate labeled with a fluor and a quencher is exposed to sample. ACE2 substrate cleavage activity frees the fluor from quenching, followed by fluorometric evaluation over time. We first established the kit’s performance characteristics on commercially purchased EDTA-anticoagulated normal human plasma. ACE2 is as Zn^2+^-requiring metalloenzyme. Since EDTA was used as the anticoagulant, to make Zn^2+^ unavailable to the Zn^2+^-requiring metalloenzymes of the enzymes involved in the coagulation cascade, we first established whether exogenously added ACE2 could be detected when added to plasma samples under our assay conditions. We found that exogenously added recombinant ACE2 could not be detected, either using commercially obtained pooled plasma or in plasma samples pooled from our patients and obtained from our sample bank, unless Zn^2+^ was added or unless the plasma was diluted (FIG 2). Since no native ACE2 activity should be detectable in the EDTA-anticoagulated plasma, any ACE2 substrate cleavage activity observed in the EDTA-anticoagulated plasma should be due to something other than ACE or another Zn^2+^ metalloprotease.

**FIG 2.**
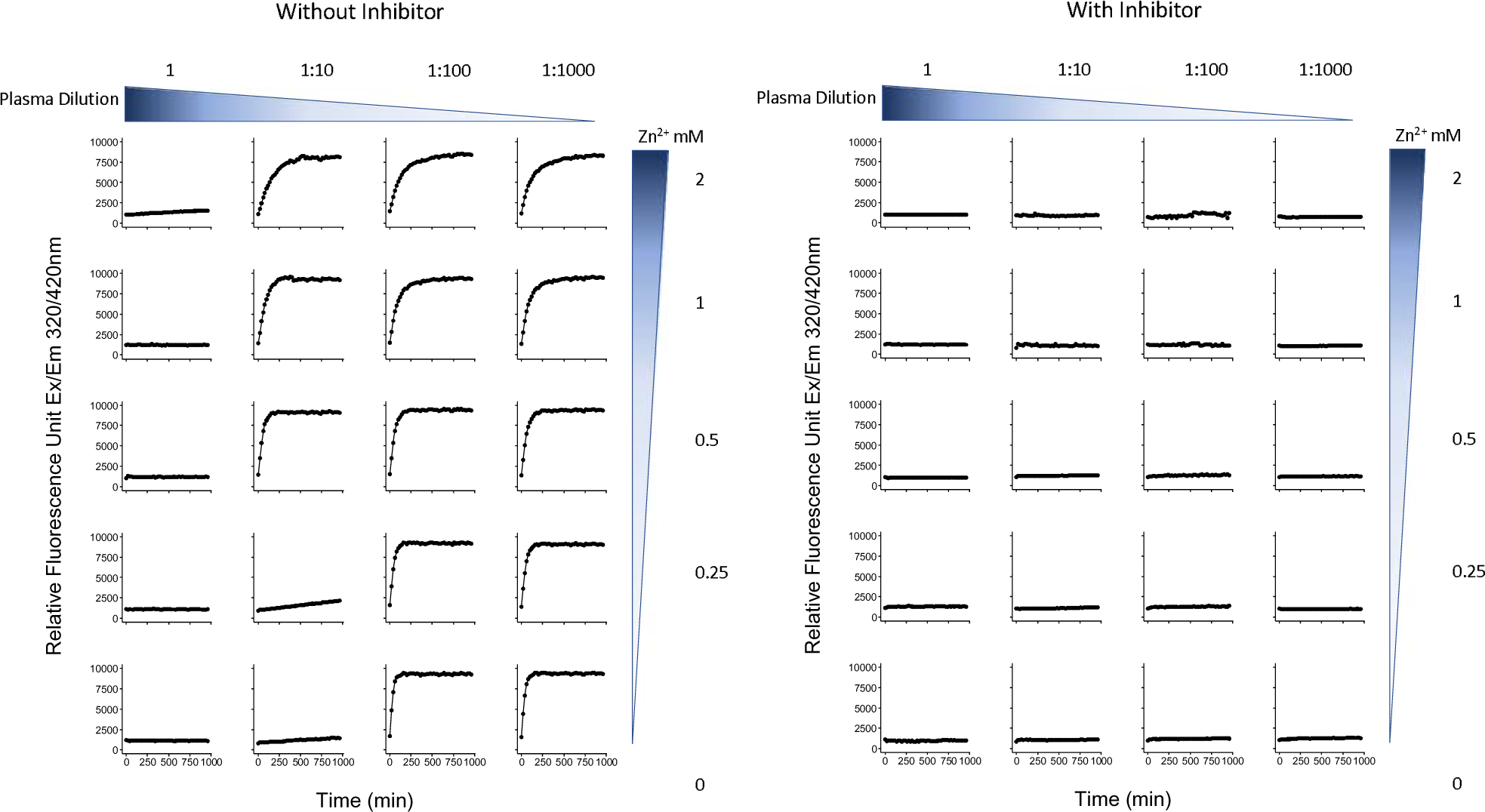
Inhibition of exogenously added recombinant ACE2 substrate cleavage activity in undiluted EDTA-anticoagulated pooled plasma samples with and without added ZnCl_2_. ACE2 is a Zn^2+^ metalloprotease. In this ACE2 spiking experiment, no ACE2 activity was detectable when exogenous ACE2 was added to the plasma, unless Zn^2+^ was also added, or unless the plasma was diluted prior to the addition of the spiked-in ACE2. Adding an ACE2 inhibitor blocked detection of the ACE2 activity in all conditions, indicating that the activity was specific for ACE2. The findings indicate that no native ACE2 activity should be detectable in the EDTA-anticoagulated plasma and so any ACE2 substrate cleavage activity observed in the EDTA-anticoagulated plasma should be due to something other than ACE or another Zn^2+^ metalloprotease.

We performed the ACE2 assay on EDTA-anticoagulated (obtained using clinical standard lavender top tubes) research participant plasma samples obtained at about the 7^th^ day of hospitalization for a cohort of 67 COVID-19 patients with moderate to severe disease. Using these samples, without dilution and without added Zn^2+^, we found that we could detect cleavage of the ACE2 peptide substrate in some of the patients, suggesting that the ACE2 peptide substrate cleavage activity was produced by an agent with co-factor characteristics different from canonical ACE2 (FIG 3). FIG 4 shows a scatter plot of the RFU values for the 67 patients studied.

**FIG 3.**
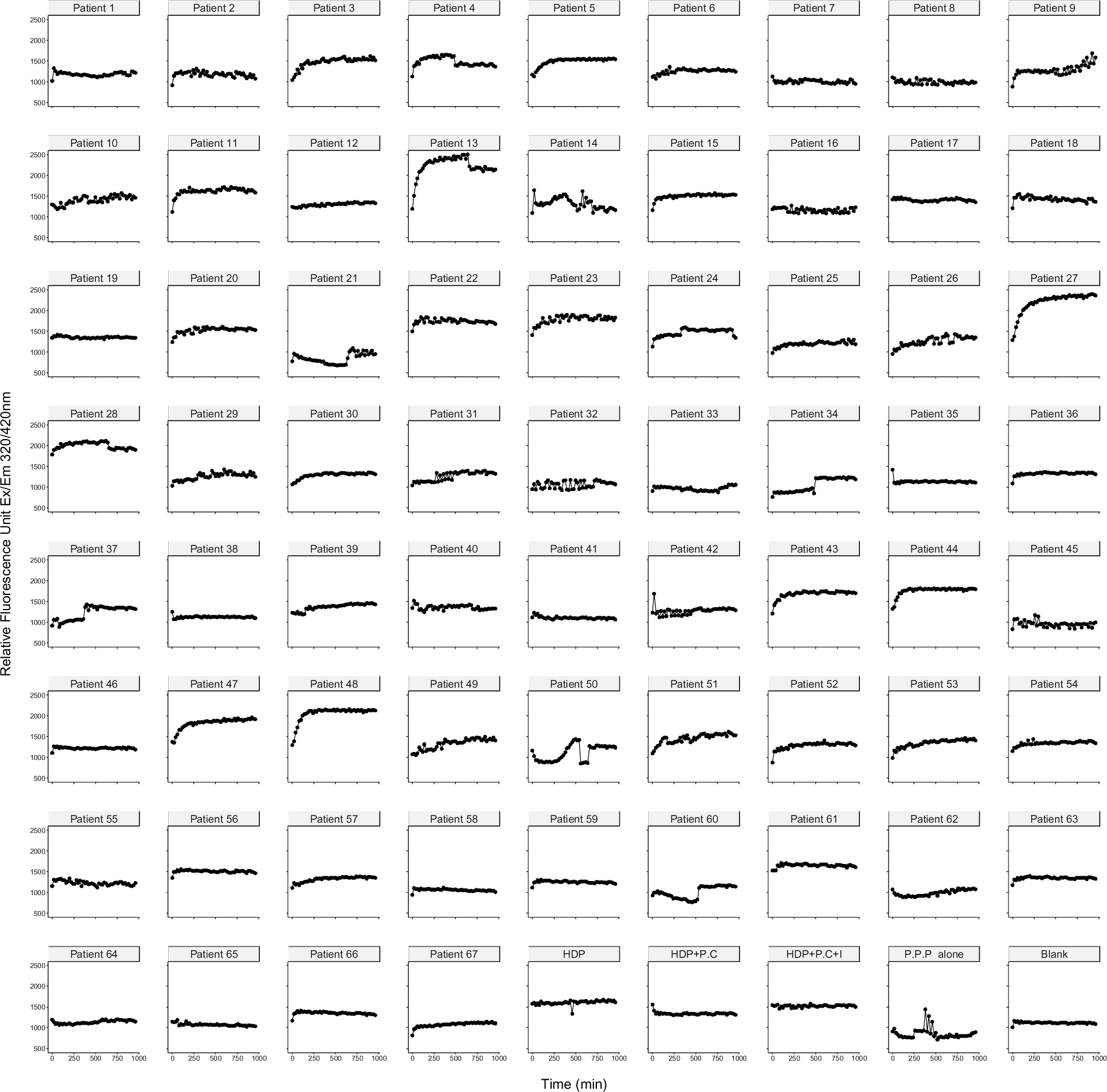
Survey of ACE2 substrate cleavage activity in plasma from 67 patients with COVID-19 sampled at about day 7 of hospitalization. The plots present the results of the fluorometric assays for ACE2 substrate cleavage activity run on the 67 patients, plus control samples. We identified plasma from 11 patients (4, 5, 9, 11, 13, 15, 27, 43, 44, 47, 48) that showed an ability to cleave ACE2 substrate. (HDP: commercially purchased pooled healthy donor plasma; HDP+P.C: HDP with added recombinant human ACE2; HDP+P.C+I: HDP+P.C.+ ACE2; inhibitor; P.P.P alone: pooled plasma from negative samples).

**FIG 4.**
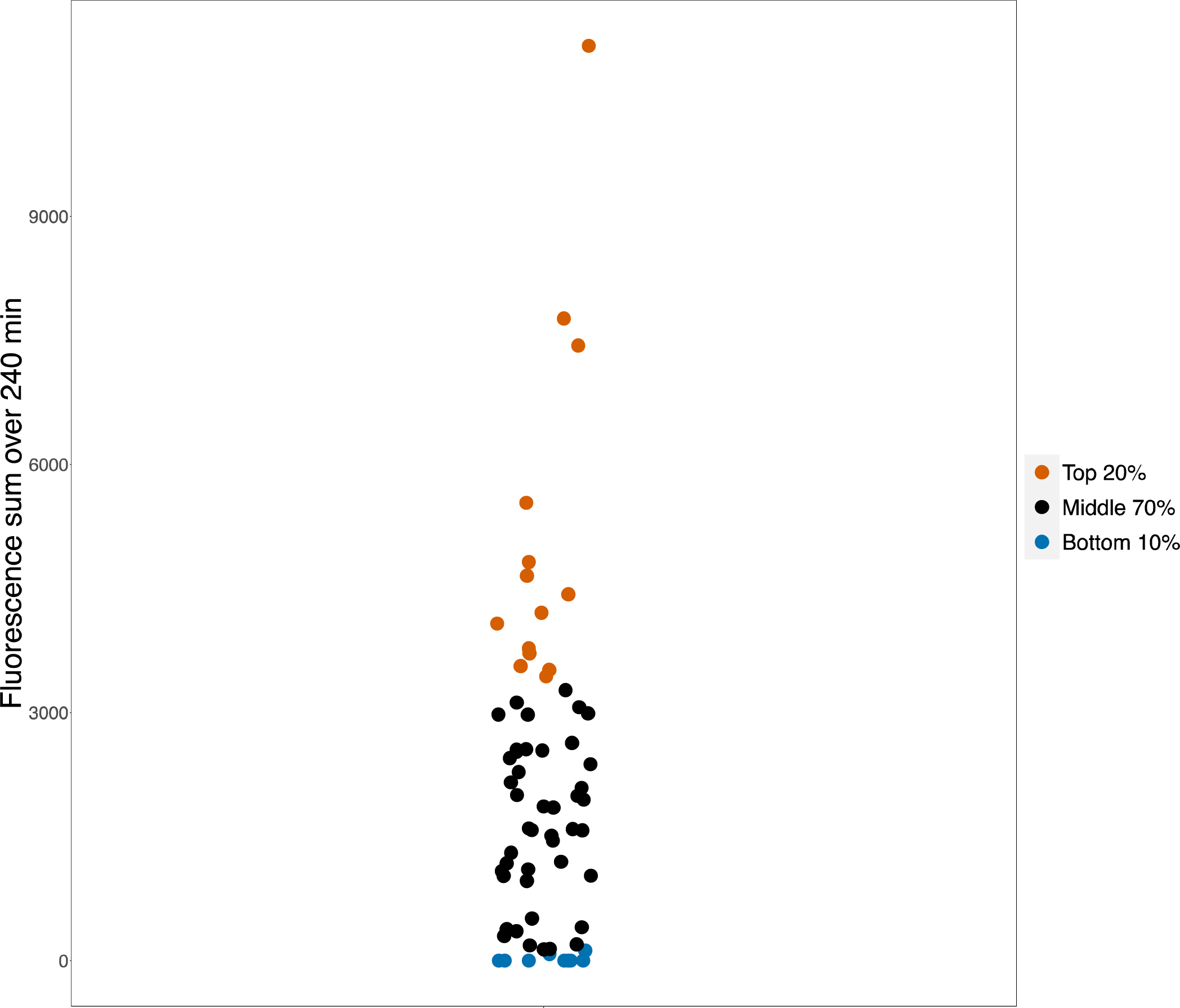
Scatter plot of the background corrected RFU values for the plasma samples from the 67 patients evaluated in this study. The top 20% of values are plotted in red, the middle 70% are plotted in black, and the bottom 10% are plotted in blue. The top 20% and bottom 10% were selected for the ACE2 substrate cleavage activity assays before and after immunoglobulin depletion shown in FIG 5.

To determine whether the ACE2 substrate cleavage activity was associated with the presence of immunoglobulin in the samples, we developed an immunoglobulin depletion protocol that includes an initial 0.45 um filtration step to remove large aggregates, followed by a 100 KDa size exclusion column chromatography step, and a subsequent staphylococcal A/G bead absorption step. To test whether patients with high RFU had ACE2 substrate cleavage activity associated with immunoglobulin, we selected samples from patients who had RFU values in the top 20% RFU values and which still had sufficient plasma samples remaining for the immunoglobulin depletion procedure and subsequent ACE2 assays. This selection yielded samples from 11 patients, 4, 5, 9, 11, 13, 15, 27, 43, 44, 47, and 48. We also selected samples from 3 patients (17, 64, 64) who were among the patients having the bottom 10% of ACE2 substrate cleavage activity and for which sufficient sample was available for the immunoglobulin depletion procedure as negative control. To determine whether our immunoglobulin depletion procedure also depleted IgM in addition to IgG, selected additional samples for which there sufficient plasma remained for the assays, patients 4, 5, 11, 13, 15, 27, 43, 44, 64, and the pooled healthy donor plasma, and determined IgM concentrations before and after immunoglobulin depletion. We also included a purchased pooled EDTA-anticoagulated plasma sample as an additional control. For both IgG and IgM, we found that our depletion procedure removed more than 99.99% of the immunoglobulins.

Table 3 show the results of the immunoglobulin depletion protocol applied to the patient plasma samples from these patients. The process depleted IgG by a factor of >>99.99%, leaving IgG in the samples at <2.5 ng/ml for all samples. Table 4 shows the results of assays for IgM run on selected samples for which sufficient sample remained for the assays. For all samples assayed, the immunoglobulin depletion procedure removed >99.9% of IgM.

**Table 3.** Plasma IgG concentration before and after antibody depletion for selected patients exhibiting ACE2 substrate cleavage activity in FIG 2, then re-assayed for ACE2 substrate cleavage activity with results shown in FIG 4.

| Patient<br>(#) | IgG Concentration |  | IgG depletion<br>efficiency<br>(%) |
| --- | --- | --- | --- |
|  | Pre<sup>a</sup><br>mg/ml | Post<sup>b</sup><br>ng/ml |  |
| 4 | 14.62 | 0.67 | >99.99% |
| 5 | 12.50 | 1.48 | >99.99% |
| 9 | 24.40 | 0.96 | >99.99% |
| 11 | 75.41 | 0.17 | >99.99% |
| 13 | 26.35 | 2.46 | >99.99% |
| 15 | 13.27 | 2.47 | >99.99% |
| 27 | 10.53 | 2.40 | >99.99% |
| 43 | 29.07 | 0.87 | >99.99% |
| 44 | 11.17 | 2.29 | >99.99% |
| 47 | 13.15 | 1.85 | >99.99% |
| 48 | 12.32 | 2.06 | >99.99% |

a: IgG concentration in plasma before antibody depletion
b: IgG concentration in plasma after antibody depletion by ultrafiltration and Protein A/G beads

**Table 4.** Plasma IgM concentration before and after antibody depletion for selected patients.

| Patient (#) | IgM Concentration (mg/ml) |  | IgM Depletion Efficiency (%) |
| --- | --- | --- | --- |
|  | Pre<sup>a</sup> | Post<sup>b</sup> |  |
| 4 | 0.875 | 1.43E-05 | >99.99% |
| 5 | 0.753 | 1.44E-05 | >99.99% |
| 11 | 0.7916 | 7.99E-06 | >99.99% |
| 13 | 0.644 | 1.41E-05 | >99.99% |
| 15 | 2.250 | 1.08E-05 | >99.99% |
| 27 | 0.549 | 1.40E-05 | >99.99% |
| 43 | 1.560 | 1.34E-05 | >99.99% |
| 44 | 3.243 | 1.41E-05 | >99.99% |
| 64 | 0.915 | 8.22E-06 | >99.99% |
| Pooled Healthy Donor | 0.4852 | 1.3568E-05 | >99.99% |

a: IgM concentration in plasma before antibody depletion
b: IgM concentration in plasma after antibody depletion by ultrafiltration and Protein A/G beads

We then re-assayed these samples to determine whether immunoglobulin depletion was associated with elimination of the ACE2 substrate cleavage activity (FIG 5), and found that for the 11 tested patients with ACE2 substrate cleavage activity in this cohort, depletion of the immunoglobulin was accompanied by elimination of ACE2 substrate cleavage activity.

**FIG 5.**
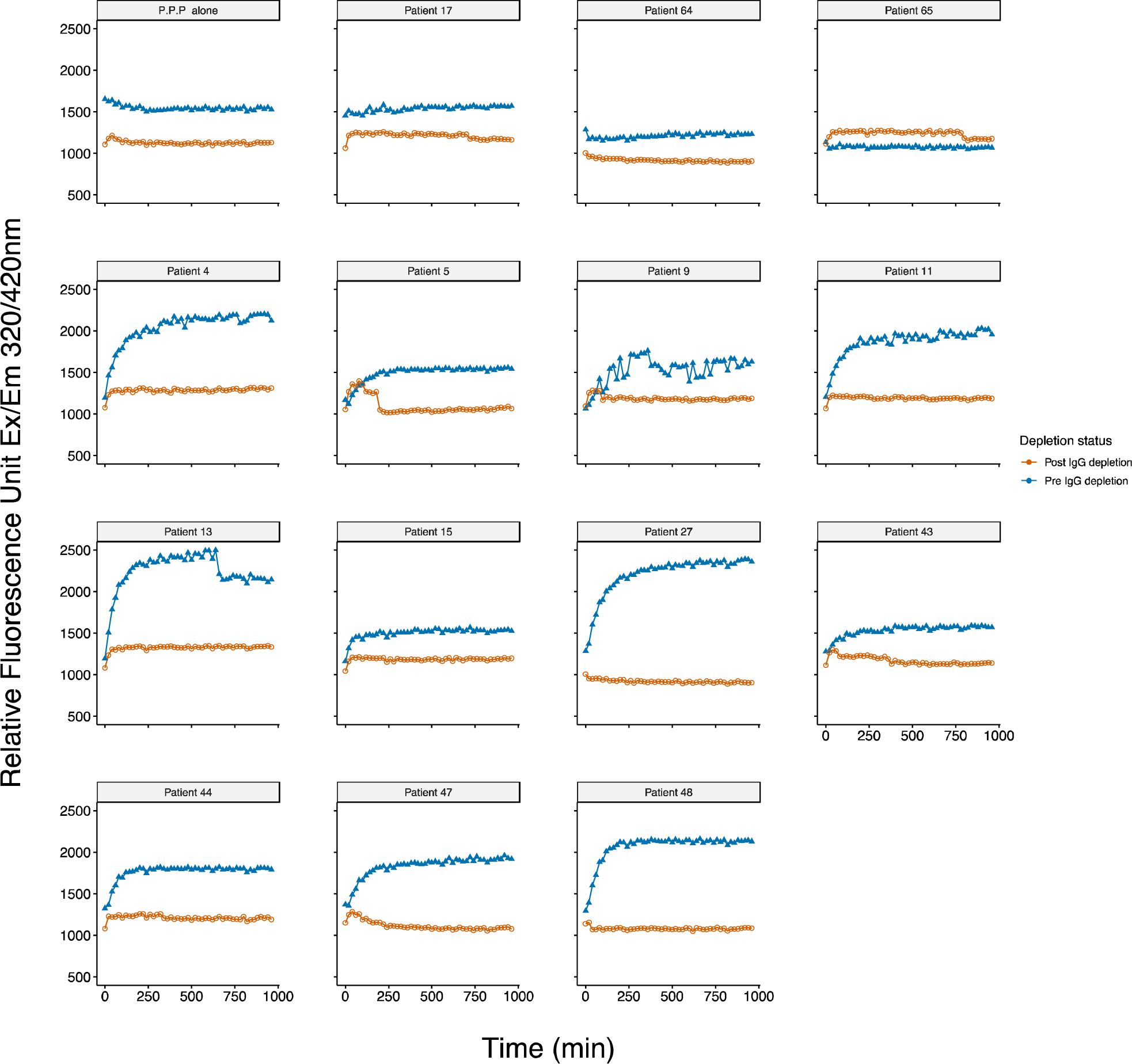
ACE2 substrate cleavage activity in patients before and after immunoglobulin depletion. The figure shows ACE2 substrate cleavage activity for 11 patients (4, 5, 9, 11, 13, 15, 27, 43, 44, 47, 48) who had among the top 20% of RFU values and for which we had sufficient plasma for immunoglobulin depletion, before (blue points) and after (red points) immunoglobulin depletion by a 100 KDa size exclusion column followed by protein A/G bead absorption. We also show ACE2 substrate cleavage activity assays before and after immunoglobulin depletion for pooled patient plasma (PPP) negative controls and samples from three arbitrarily selected patients, 17, 64, 65, that had among the bottom 10% of RFU values and had sufficient plasma remaining for the immunoglobulin depletion protocol. The data suggest that for the 11 patients demonstrating the highest ACE2 substrate cleavage activity in this cohort, the ACE2 substrate cleavage activity was depleted in an immunoglobulin-associated manner.

To determine whether the cleavage activity was specific, we used a control substrate, OMNIMMP® fluorogenic control (Enzo Life Sciences, Cat. #MML-P127), compared with ACE2 standard substrate, Mca-APK(Dnp), provided by the assay kit according to a previous study^36^. Our pre-optimization study indicated 25µM of control substate had similar background level with standard substrate when incubated with purified recombinant hACE2 positive control provided by the assay kit. Plasma samples with ACE2-like activity were incubated with Mca-APK(Dnp) as well as 25µM OMNIMMP® fluorogenic control substrate and fluorescence were read every 20min for 16 hours. The plasma samples with ACE2-like activity when incubated with Mca-APK(Dnp), had no activity when assayed with the OMNIMMP control substrate (FIG 6). A consensus sequence of Pro-X-Pro-hydrophobic/basic for the protease specificity of ACE2 has been defined ^43, 44^. Unlike Mca-YVADAPK(Dnp), a previously used substrate for caspase-1/interleukin-converting enzyme (ICE) and ACE-2 ^44, 45^, Mca-APK(Dnp) is not cleaved by caspases and Mca fluorescence is quenched by the Dnp group until cleavage (at Pro-Lys) separates them. OMNIMMP® fluorogenic substrate, Mca-Pro-Leu-OH, is used as fluorogenic Mca control peptide for Mca-APK(Dnp) substrate by the manufacturer. Studies also demonstrated that peptide substrates with similar sequences could not be hydrolyzed by human ACE2 ^43, 46^. This indicates that the catalytic activities of those samples were specific ACE2-like activities. We also incubated samples with a synthetic SARS-CoV-2 spike RBD peptide pool before adding the substrate to determine if the RBD peptide pool would compete with the ACE2 substrate. We conducted 5-fold serial dilutions of the peptide pools, starting at 10 µg per peptide, incubating the peptide competitors with plasma samples and ACE2 standard substrate, Mca-APK(Dnp). The ability to cleave the ACE2 substrate were inhibited by RBD peptides in a dose dependent manner (FIG 7 and Supplementary FIG 1) which suggested that the tiled peptides covering the RBD region of SARS-CoV-2 spike competitively inhibits the ACE2-like activities in plasma, further confirming the specificity of the ACE2-like activities in plasma.

**FIG 6.**
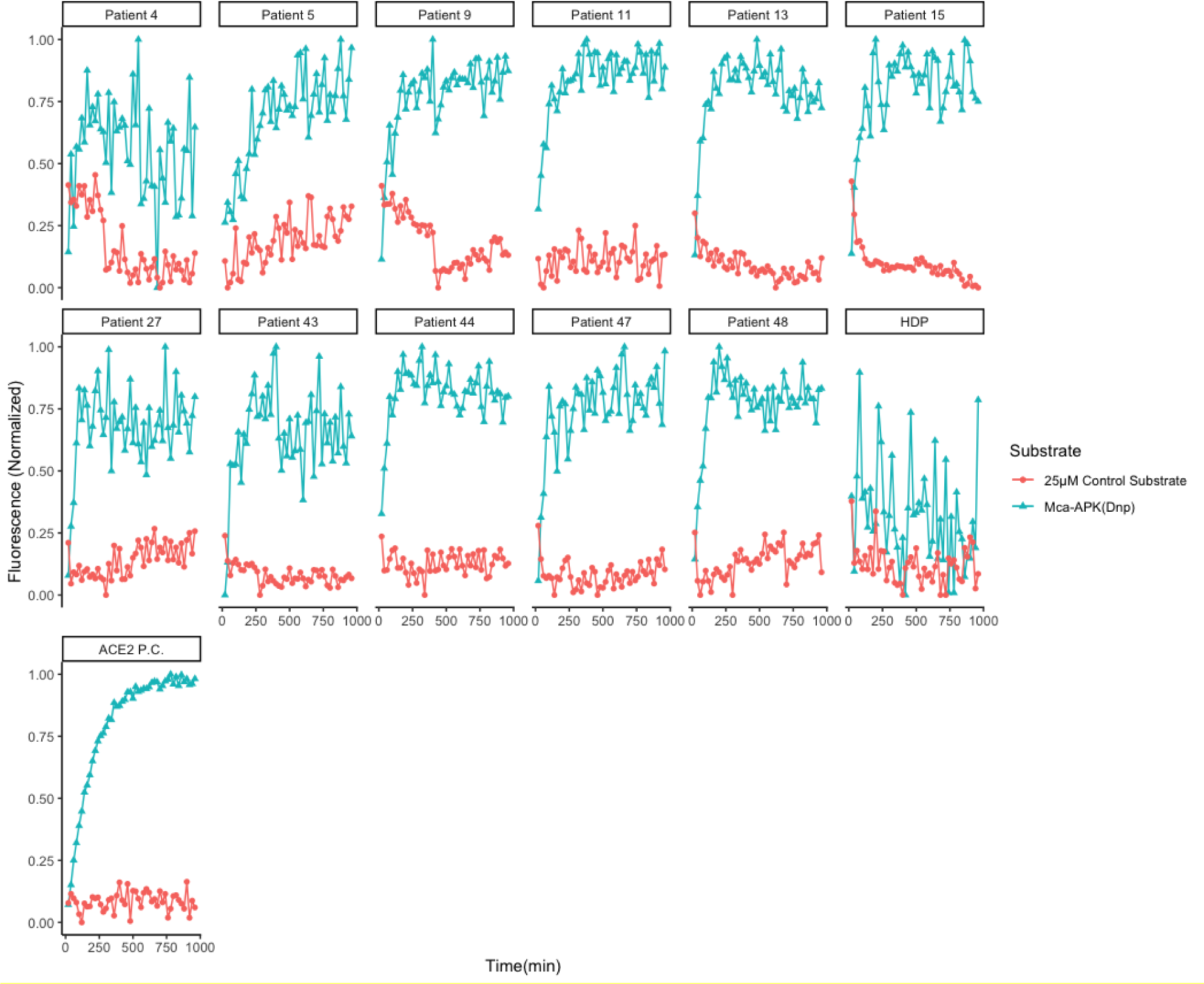
Substate cleavage specificity of patient plasma. ACE2 substrate cleavage activity specificities for 11 patients (4, 5, 9, 11, 13, 15, 27, 43, 44, 47, 48) who had among the top 20% of RFU values were measured with ACE2 standard substrate (Mca-APK(Dnp)) and control substrate (OMNIMMP® fluorogenic control). All 11 samples as well as positive control (ACE2 P.C.), purified recombinant human ACE2 provided by the assay kit, had specific cleavage activities on Mca-APK(Dnp), but no activities on control substrate. HDP: commercially purchased pooled healthy donor plasma as ACE2-activity negative control sample.

**FIG 7.**
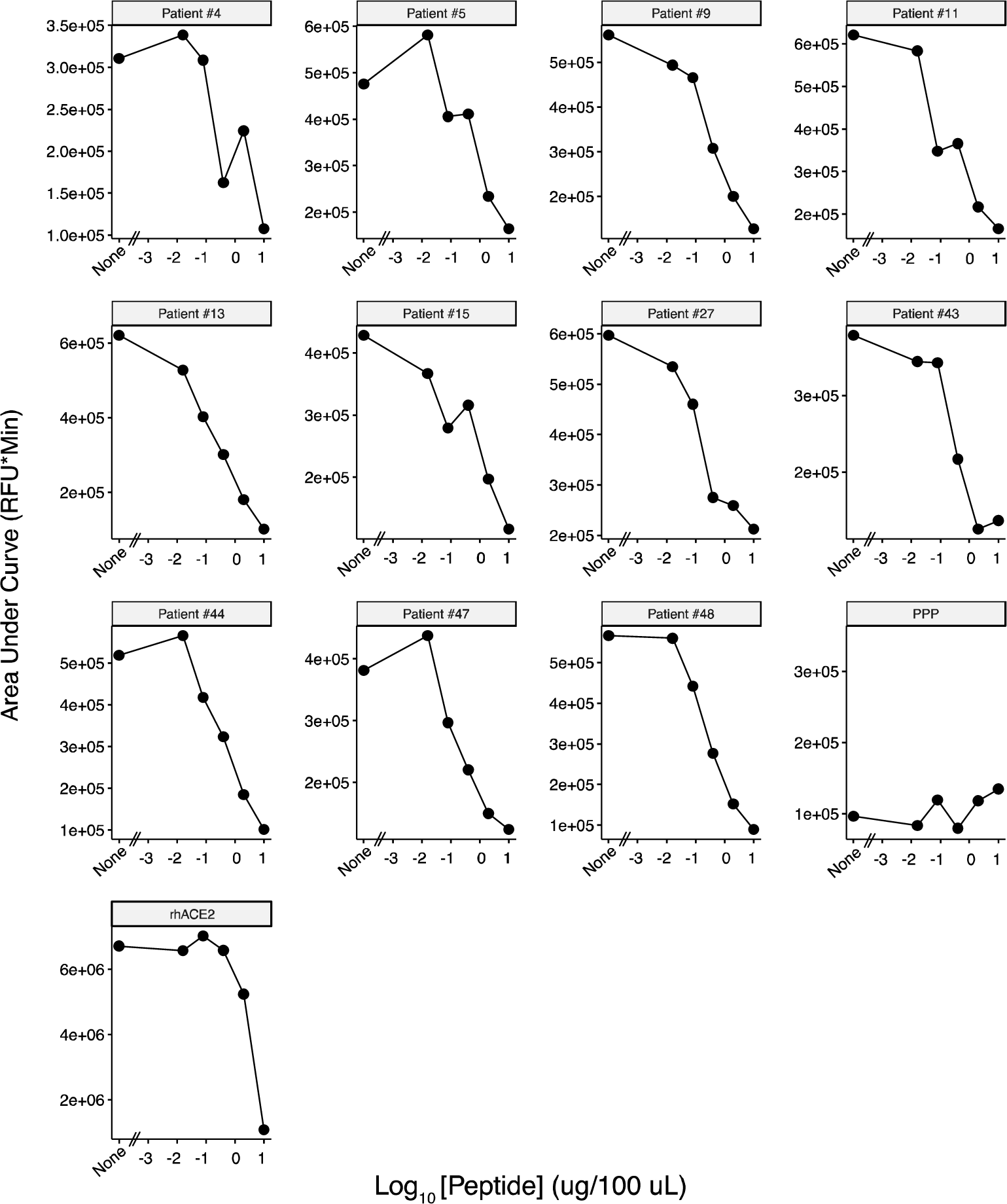
Plasma ACE2-like activity competitive inhibition by SARS-CoV-2 spike RBD peptides. ACE2 substrate cleavage activity for 11 patients (4, 5, 9, 11, 13, 15, 27, 43, 44, 47, 48) who had among the top 20% of RFU values were measured by incubation with serial diluted SARS-CoV-2 spike RBD peptide pools. ACE2-like catalytic activities in plasma were competitively inhibited by RBD peptide pool in a dose dependent manner. AUCs of each curve from samples with individual peptide amount were calculated based on all RFU readouts. rhACE2: purified recombinant human ACE2 provided by the assay kit as positive control. PPP: commercially purchased pooled healthy donor plasma as ACE2-activity negative control sample.

If our hypothesis is correct and the ability to cleave an ACE2 substrate is related to Abs against the RBD, then we should be able to observe a correlation between ACE2 substrate cleavage activity and the amount of Ab directed against the RBD in the patients’ samples. We assayed for anti-RBD Ab binding activity using a modification of an ImmunoCAP^TM^ Assay activity using a Phadia 250 instrument and S-RBD (FIG 8). We found that there was a significant correlation between the amount of Ab in the patient plasma samples capable of binding the RBD and the ACE2 substrate cleavage activity, suggesting that the ACE2 cleavage activity was related to the amount of anti-RBD Ab present in the plasma sample.

**FIG 8.**
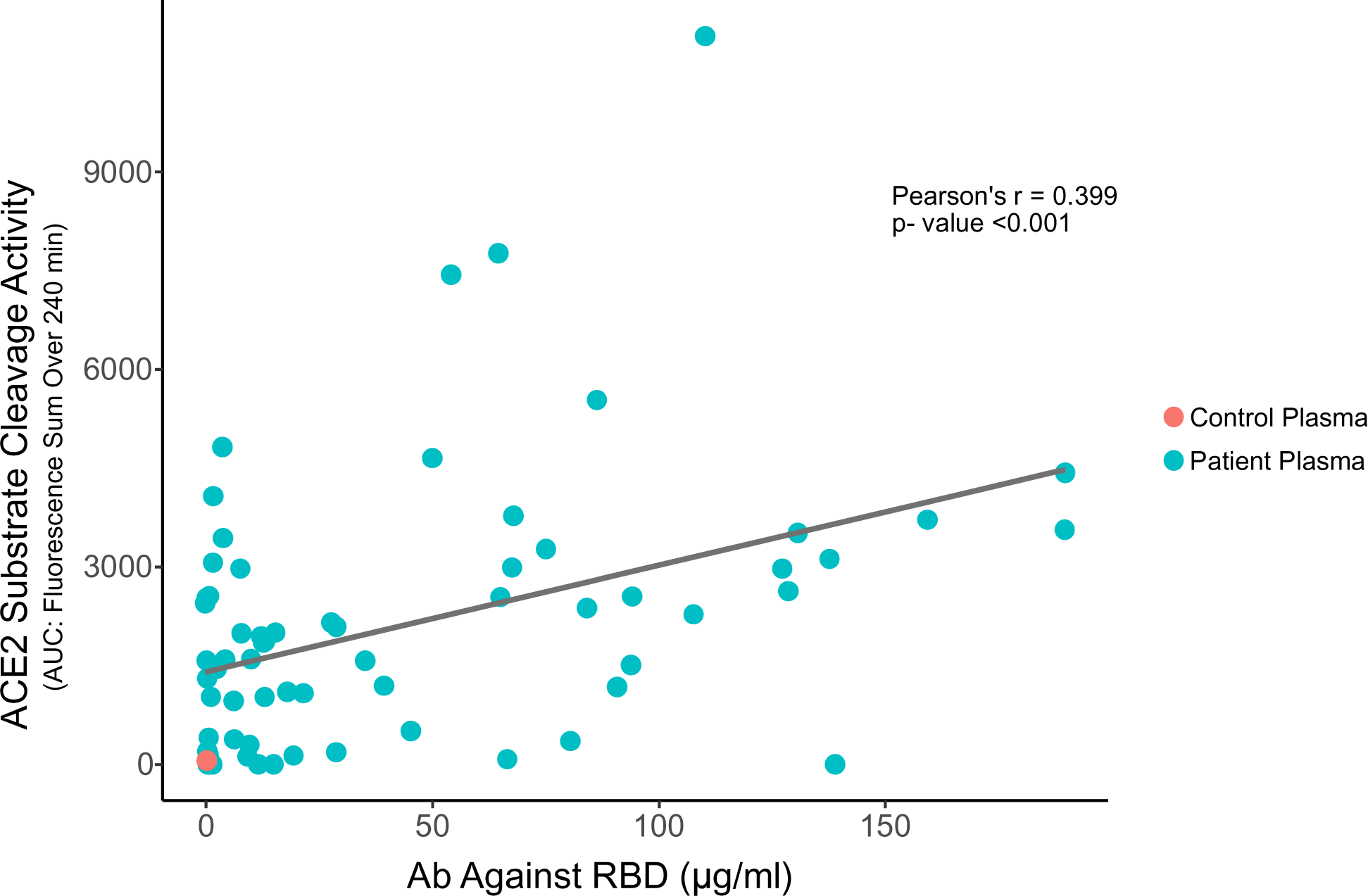
Correlation of ACE2 substrate cleavage activity (ACE2 Activity) with anti-RBD antibody (Ab Against RBD) in patient plasma. Anti-RBD antibody concentrations in plasma samples were determined using ImmunoCAP^TM^ antibody binding assay. The negative control pooled human plasma is indicated by the red symbol. The patient plasma sample are indicated in green. The negative control plasma values were omitted in the calculation of the r-and p-values.

## Discussion

Our experiments indicate that a fraction of patients with COVID-19 have an activity in plasma capable of cleaving a substrate for ACE2. The ACE2 substrate cleavage activity has divalent cation requirements different from native ACE2 and collocates with immunoglobulin presence during an immunoglobulin depletion procedure, suggesting that these patients may have antibodies with abzyme activity.

The ACE2 substrate cleavage activity showed a significant correlation with Ab directed against the RBD, further supporting the hypothesis that some anti-RBD Abs have ACE2 substrate cleavage activity, but there was considerable spread in the identification of ACE2 substrate cleavage activity observed for different levels of anti-RBD Ab. However, this spread would be expected if only a small fraction of Abs elicited against the RBD in fact had ACE2 substrate cleavage activity, with the initiation and maturation of Ab responses directed against the RBD resulting, in part, from non-specific processes.

Since abzymes can have a broadened substrate specificity compared to the original enzyme that they mirror, it is possible that abzymes made in response to SARS-COV-2 infection may cleave a range of substrates, including substrates that form components of important regulatory cascades, like clotting, blood pressure regulation, and inflammation processes, which could account for aspects of COVID-19 that cannot be directly attributed to viral infection and cell and tissue destruction.

Our study had several strengths. We studied a moderate number of patients, with samples collected prospectively from acutely hospitalized patients with clearly established COVID-19 relatively early in their disease course, but likely approaching at least 2 weeks following initial infection. The patients almost certainly were not admitted immediately following initial infection and the samples were obtained about a week after admission, providing enough time for the development of an initial antibody response to SARS-CoV-2 infection. We used a standard, widely-employed kit for ACE2 peptide substrate cleavage activity ^36, 44, 47^ of which the ACE2 standard substrate, Mca-APK(Dnp), was previously developed as an improved fluorescence substrate with more efficient intramolecular fluorescence quenching and enhanced fluorescence increase over background after complete hydrolysis compared with other peptide substates^44^, standard size exclusion columns, protein A/G bead absorption immunoglobulin depletion techniques, and assays for immunoglobulins. The data showing that the ACE2 substrate cleavage activity was present even when there was sufficient EDTA to chelate the Zn^2+^ essential for ACE2 activity suggests that the ACE2 substrate cleavage activity had important characteristics different from native ACE2. The catalytic activities in plasma were ACE2 substrate specific and were competitively inhibited by peptides covering the SARS-CoV-2 spike RBD, further indicating that the cleavage activity was specific. The data that the ACE2 substrate cleavage activity was removed from the samples following processing with size exclusion columns and absorption with protein A/G beads suggests that the non-Zn^2+^-requiring ACE2 substrate cleavage activity co-localizes with immunoglobulin and the data that ACE2 substrate cleavage activity correlated with Ab against the RBD support the hypothesis that the patients with ACE2 substrate cleavage activity may have developed abzymes with this activity.

However, our study has several limitations. We only studied a limited subset of the patients admitted to our hospital, from patients who consented to the sample banking and study and for whom sufficient sample volumes were available. A contemporaneously collected, matched, prospective set of samples from patients without COVID-19 would have been ideal negative controls, but samples from such subjects were not obtained as part of the sample banking effort. We also only conducted the assays on patient samples obtained at a single time, about 7 days after hospitalization, so we have no data from patients who were discharged, transferred, or died prior to day 7 of hospitalization. For this study, we were unable to correlate the presence of ACE-2 abyme activity with clinical disease features. More importantly, in this study we have no data from patients later in their disease course or following convalescence from the acute disease. Inflammatory, coagulopathic, and physiologic dysregulatory phenomena predominate in the later stages of COVID-19 and at least some aspects of PASC and MIS-C appear to result from immune-mediated phenomena. However, some of these patients are treated with anti-inflammatory agents, including intravenous immunoglobulin, which would make the study of these patients for abzyme-like activity difficult. If abzymes with promiscuous substrate specificities develop in some COVID-19 patients, some of the clinical features of those disorders could potentially be attributed to abzymes, but this study is not able to directly address such additional hypotheses, which would require additional work. In addition, since abzymes can show substantial substrate specificity spread, it is possible that additional patients in our cohort had abzymes with substrate specificity sufficiently divergent from angiotensinogen that it was not detectable using our assay. It is also possible that COVID-19 patients develop anti-RBD antibodies with abzyme-like activity, but that these antibodies have no clinical significance. Some fraction of normal individuals may have ACE2-like abzyme activity, but this activity could have been diluted in the purchased pooled normal control plasma. However, if the abzyme activity in such patients was similar to that observed in the individual positive patients in this study, that activity would still be ∼15% of the activity of the positive patients, given the prevalence of detectable abzyme activity in our cohort, and so would have been likely detectable.

Finally, our study is merely associational. It is possible that we depleted some other, non-immunoglobulin component of COVID-19 patient plasma that has the ability to cleave the ACE2 substrate peptide with our 100 kDa size exclusion plus protein A/G treatment procedure. To unequivocally establish that some COVID-19 patients develop anti-SARS-CoV-2 immune responses that include abzymes, it would be necessary to isolate discrete monoclonal antibodies from patients with ACE2 substrate cleavage activity, and then characterize those monoclonal antibodies in detail. Such work lies outside the perimeters of this current study. Additional studies of potential abzyme-like activity in COVID-19 patients with different clinical disease manifestations and at different points in their disease course, including long after acute COVID-19, would be needed to establish the overall significance of abyzmes in COVID-19 disease. While understanding the detailed pathogenic significance of abzymes in COVID-19 still requires substantial investigation, the existence of abzymes in COVID-19 is increasingly well-established. In addition to the studies reported here, McConnell and co-workers ^48^ recently described the existence of abzymes in convalescent plasma COVID-19 patients capable of cleaving spike protein with virus neutralization. That two different groups independently found evidence of abzyme activity in plasma from COVID-19 patients provides significant support for the hypothesis that COVID-19 patients can develop antibodies with proteolytic activity.

Nevertheless, our study offers potential additional insight into some of the difficult-to-understand clinical features of COVID-19, and if some of the difficult manifestations of COVID-19 are related to the induction of antibodies with abzyme-like activity, that finding may suggest new therapeutic interventions. Some other viruses also have also evolved to use enzymes located on the target host cell surface as their receptors, so it is conceivable that those other viral infections may also elicit clinically problematic anti-viral antibodies. The number of spike protein RBDs presented to the immune system in a natural infection is vastly greater than the number of RBDs presented to the immune system following vaccination, offering many more opportunities for the induction of wide-ranging antibody responses, unlike the antigen exposure resulting from vaccination. While complications following vaccination are very rare and the risk-benefit calculation greatly favors vaccination, some of the adverse side effects of SARS-CoV-2 vaccines might also be attributable to the rare induction of anti-RBD antibodies with catalytic activity.

## Institutional Review Board Statement

The sample collection protocol was approved by the UVA IRB (HSR #200110), and approval was obtained to work on the specimens (HSR #HSR200362).

## Informed Consent Statement

Informed consent was obtained from all participants involved in the study.

## Data Availability Statement

Data is presented in the manuscript and the supplements. Raw data can be obtained on request.

## Data Availability

All data produced in the present work are contained in the manuscript and are available upon reasonable request.

## Acknowledgments and Funding

The study was supported by internal funding from the University of Virginia, including the Manning Fund for COVID-19 Research at UVA the Ivy Foundation, the Pendleton Laboratory Fund for Pediatric Infectious Disease Research, a College Council Minera Research Grant, the Coulter Foundation, and NIAID, NIH (R01 AI176515) with sample acquisition costs supported by the HHV-6 Foundation. We thank the study participants for graciously agreeing to be research participants, and the research support staff who enrolled the study participants (led by Linda Bailes RN, Allison Raymond RN and Sarah Struchen RN). We also thank the University of Virginia Biorepository and Tissue Research Facility for processing the samples used in the study and for providing specimens (Technical Director, Patcharin Pramoonjago PhD). We also want to appreciate Dr. John Lazo, Dr. Elizabeth Sharlow, Dr. Kevin Lynch, and Stefan Hargett from Department of Pharmacology, University of Virginia for their great help on instruments for the assays.

## Supplementary Data Legends

S1. Supplementary Figure 1. Plasma ACE2-like activity competitive inhibition by SARS-CoV-2 spike RBD peptides. A. ACE2 substrate cleavage activity for 11 patients (4, 5, 9, 11, 13, 15, 27, 43, 44, 47, 48) who had among the top 20% of RFU values were measured by incubation with serial diluted SARS-CoV-2 spike RBD peptide pools. ACE2-like catalytic activities in plasma were competitively inhibited by RBD peptide pool in a dose dependent manner. B. ACE2 activity of purified recombinant human ACE2 provided by the assay kit as positive control incubated with serial diluted SARS-CoV-2 spike RBD peptide pools. PPP: commercially purchased pooled healthy donor plasma as ACE2-activity negative control sample.

