## Supplementary material for "ACE-2-like enzymatic activity is associated with immunoglobulin in COVID-19 patients": FIG 7 and Supplementary FIG 1

Supplementary Figure 1. Plasma ACE2-like activity competitive inhibition by SARS-CoV-2 spike RBD peptides. A. ACE2 substrate cleavage activity for 11 patients (4, 5, 9, 11, 13, 15, 27, 43, 44, 47, 48) who had among the top 20% of RFU values were measured by incubation with serial diluted SARS-CoV-2 spike RBD peptide pools. ACE2-like catalytic activities in plasma were competitively inhibited by RBD peptide pool in a dose dependent manner. B. ACE2 activity of purified recombinant human ACE2 provided by the assay kit as positive control incubated with serial diluted SARS-CoV-2 spike RBD peptide pools. HDP: commercially purchased pooled healthy donor plasma as ACE2-activity negative control sample.

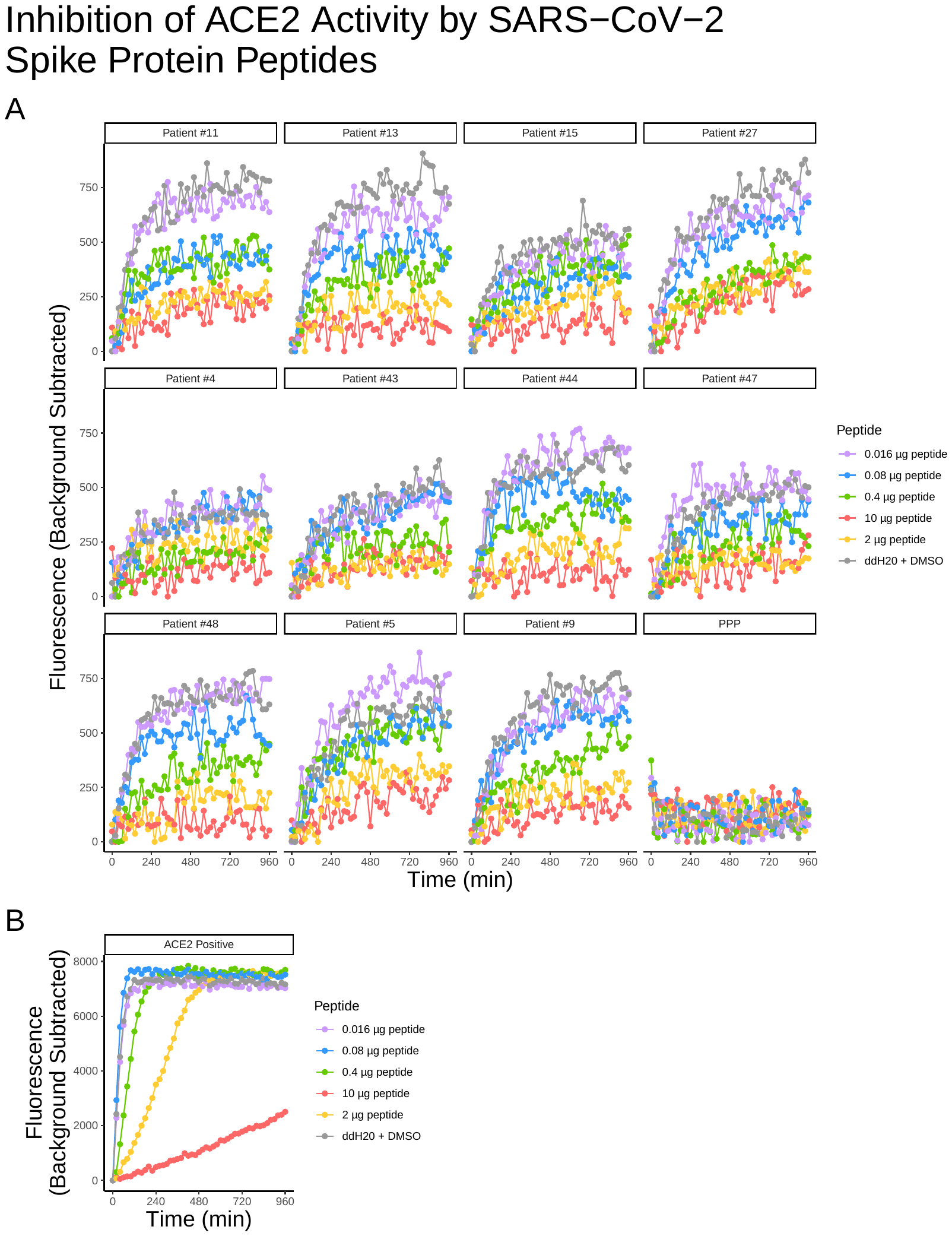
